# Evaluating the effect of rapamycin treatment in Alzheimer's Disease and aging using *in vivo* imaging: the ERAP phase IIa clinical study protocol

**DOI:** 10.1101/2024.02.19.24302922

**Authors:** Jonas E. Svensson, Martin Bolin, Daniel Thor, Pete A. Williams, Rune Brautaset, Marcus Carlsson, Peder Sörensson, David Marlevi, Rubens Spin-Neto, Monika Probst, Göran Hagman, Anton Forsberg Morén, Miia Kivipelto, Pontus Plavén-Sigray

**Affiliations:** Centre for Psychiatry Research, Department of Clinical Neuroscience, Karolinska Institutet and Stockholm Health Care Services, Region Stockholm, Stockholm, Sweden; Theme Inflammation and Aging, Karolinska University Hospital, Stockholm, Sweden; Department of Medical Radiation Physics and Nuclear Medicine, Karolinska University Hospital, Stockholm, Sweden; Department of Oncology-Pathology, Karolinska Institutet, Stockholm, Sweden; Department of Clinical Neuroscience, Division of Eye and Vision, St. Erik Eye Hospital, Karolinska Institutet, Stockholm, Sweden; Clinical Physiology, Department of Clinical Sciences Lund, Lund University, Skåne University Hospital, Lund, Sweden; Department of Clinical Physiology, Karolinska University Hospital, and Karolinska Institutet, Stockholm, Sweden; Department of Medicine Solna, Karolinska Institutet, Stockholm, Sweden; Institute for Medical Engineering and Science, Massachusetts Institute of Technology, Cambridge, Massachusetts, USA; Department of Dentistry and Oral Health, Section for Oral Radiology, Aarhus University, Aarhus C, Denmark; Department of Diagnostic and Interventional Neuroradiology, Klinikum rechts der Isar, School of Medicine, Technical University of Munich, Munich, Germany; Division of Clinical Geriatrics, Department of Neurobiology, Care Sciences, and Society, Karolinska Institutet, Stockholm, Sweden; Ageing Epidemiology Research Unit (AGE), School of Public Health, Faculty of Medicine, Imperial College London, UK; Institute of Public Health and Clinical Nutrition, University of Eastern Finland, Kuopio, Finland; Neurobiology Research Unit, Copenhagen University Hospital, Rigshospitalet, Copenhagen, Denmark

**Keywords:** Rapamycin, Sirolimus, Alzheimer’s disease, clinical trial, PET, MRI, CT, OCT, geroprotection, aging

## Abstract

**Background:** Rapamycin is an inhibitor of the mechanistic target of rapamycin (mTOR) protein kinase, and pre-clinical data demonstrate that it is a promising candidate for a general gero- and neuroprotective treatment in humans. Results from mouse models of Alzheimer’s disease have shown beneficial effects of rapamycin including preventing or reversing cognitive deficits, reducing amyloid oligomers and tauopathies and normalizing synaptic plasticity and cerebral glucose uptake. The “Evaluating rapamycin treatment in Alzheimer’s disease using positron emission tomography” (ERAP) trial aims to test if these results translate to humans through evaluating the change in cerebral glucose uptake following six months of rapamycin treatment in participants with early-stage Alzheimer’s disease.

**Methods:** ERAP is a six month long, single-arm, open-label, phase IIa biomarker driven study evaluating if the drug rapamycin can be repurposed to treat Alzheimer’s disease. Fifteen patients will be included and treated with a weekly dose of 7 mg rapamycin for six months. The primary endpoint will be change in cerebral glucose uptake, measured using [^18^F]FDG positron emission tomography. Secondary endpoints will be change in cognitive measures, markers in cerebrospinal fluid as well as cerebral blood flow measured using magnetic resonance imaging. As exploratory outcomes, the study will assess change in multiple age-related pathological processes, such as periodontal inflammation, retinal degeneration, bone mineral density loss, atherosclerosis and decreased cardiac function.

**Discussion:** The ERAP study is a clinical trial using *in vivo* imaging biomarkers to assess the repurposing of rapamycin for the treatment of Alzheimer’s disease. If successful, the study would provide a strong rationale for large-scale evaluation of mTOR-inhibitors as a potential disease modifying treatment in Alzheimer’s disease.

**Trial Registration:** ClinicalTrials.gov ID NCT06022068, date of registration 2023-08-30

## BACKGROUND

For many decades, the “amyloid hypothesis” has been the dominating scientific lead in understanding and treating Alzheimer’s disease (AD). Clinical trials that directly target amyloid plaques (such as amyloid antibodies) have however resulted in mixed success (1). Only recently have two amyloid antibodies been given accelerated approval by the FDA. The drugs are prohibitively priced and questions about their efficacy and safety profile remain (2). It is therefore crucial to explore new scientific approaches to find an efficient disease-modifying intervention. One such approach is to focus on the single largest risk factor for AD: advancing age.

It is estimated that the risk of developing AD doubles every five years over the age of 65 (3), and the risk of death from AD increases by about 700 times between the ages of 55 and 85 (4). Within the field of geroscience, which focuses on the biology of aging, an increasing number of interventions have been shown to enhance the lifespan of model organisms and slow down or prevent age-related pathology (5). One promising approach to understand and treat age-related diseases like AD is to study the effects of such interventions; defined as “geroprotective compounds”, in humans (6). Pre-clinical data suggest that the drug rapamycin is a promising candidate for this purpose (6,7).

Rapamycin, also known as *sirolimus*, is an immunosuppressive drug which has been in clinical use for more than two decades. In mice, treatment with rapamycin increases average lifespan by 10 to 15% (8). The drug has also been shown to increase healthspan in model organisms, by delaying the onset of age-related diseases (9). For example, preclinical data support a beneficial effect of rapamycin (or it’s analogous) on periodontitis(10), retinal pathologies (11,12), atherosclerosis (13,14); cardiac dysfunction (15,16), and bone mass loss (17,18). Such diseases are commonly manifested with increasing age and are considered frequent comorbidities to AD (19–31).

There is a large body of preclinical data suggesting that repurposing rapamycin to treat AD could be effective (6,7). In several independent mice models of AD, rapamycin has been shown to prevent and reverse cognitive deficits (32,33), reduce amyloid oligomers and tauopathies (34,35), normalize synaptic plasticity (36), cerebral glucose uptake and (33) vascular cognitive impairment (37). Additionally, in transgenic rodent models of AD, rapamycin has demonstrated neuroprotective effects, by restoring blood-brain barrier function (32) and improving neurovascular coupling (38).

Despite promising preclinical data supporting rapamycin as an effective agent in alleviating or reversing AD pathology, no large-scale human clinical studies have been initiated. Currently, only one phase II trial is ongoing (ClinicalTrials.gov ID: NCT04629495).

Conducting randomized controlled trials (RCTs) with symptom ratings (such as cognitive ability) as endpoints is challenging due to the need for large sample sizes and high costs. An alternative approach is to assess the impact of candidate interventions on AD biomarkers before initiating such large-scale RCTs. By focusing on well-established and precise biomarkers of the disease, rather than symptom ratings, evidence of slowing or even reversal of pathology can be obtained with much smaller sample sizes (39,40).

The purpose of the study “Evaluating rapamycin treatment in Alzheimer’s disease using positron emission tomography” (ERAP) is to assess the effect of rapamycin in treating early-stage AD. This will be done by measuring change in biomarkers using *in vivo* imaging modalities, such as positron emission tomography (PET) and magnetic resonance imaging (MRI), as well as biomarker changes in cerebrospinal fluid (CSF). We will test the hypothesis that rapamycin can reverse AD-associated brain pathologies, resulting in, primarily, an increase in neuronal glucose metabolism, and secondarily, an improved cerebral blood flow and a decrease in tau and amyloid protein aggregates in the CSF. We will also record the occurrence of adverse events and investigate pharmacokinetic properties of the drug. Further, we aim to explore the effect of rapamycin on other age-related pathologies in the body, using different imaging techniques to assess change in i) periodontal inflammation, ii) retinal structures, iii) bone mineral density, iv) atherosclerosis, as well as v) cardiac function. The results from this phase IIa trial will be used to inform on the feasibility of conducting a larger controlled trial in the future.

## METHODS

### Study Design

ERAP is a single-centre, open-label, one-arm, phase IIa intervention study. Fifteen patients diagnosed with early-stage AD will be recruited from the Karolinska University Hospital, Medical Unit Aging Memory clinic, located in Solna, Stockholm, Sweden. The unit is a specialized outpatient clinic that examines individuals referred by general practitioners in primary and occupational health care in the northern catchment of Stockholm, as well as individuals younger than 70 years in the entire Stockholm region (41).

Following baseline measurements, all participants will receive a weekly oral dose of 7 mg rapamycin (Tablet Rapamune®) for a duration of six months. Throughout the study, participants will be continuously monitored for safety and adverse events. By the end of the treatment period, follow-up measurements will be conducted. Figure 1 presents a schematic overview of the study timeline for each participant.

**Figure 1.**
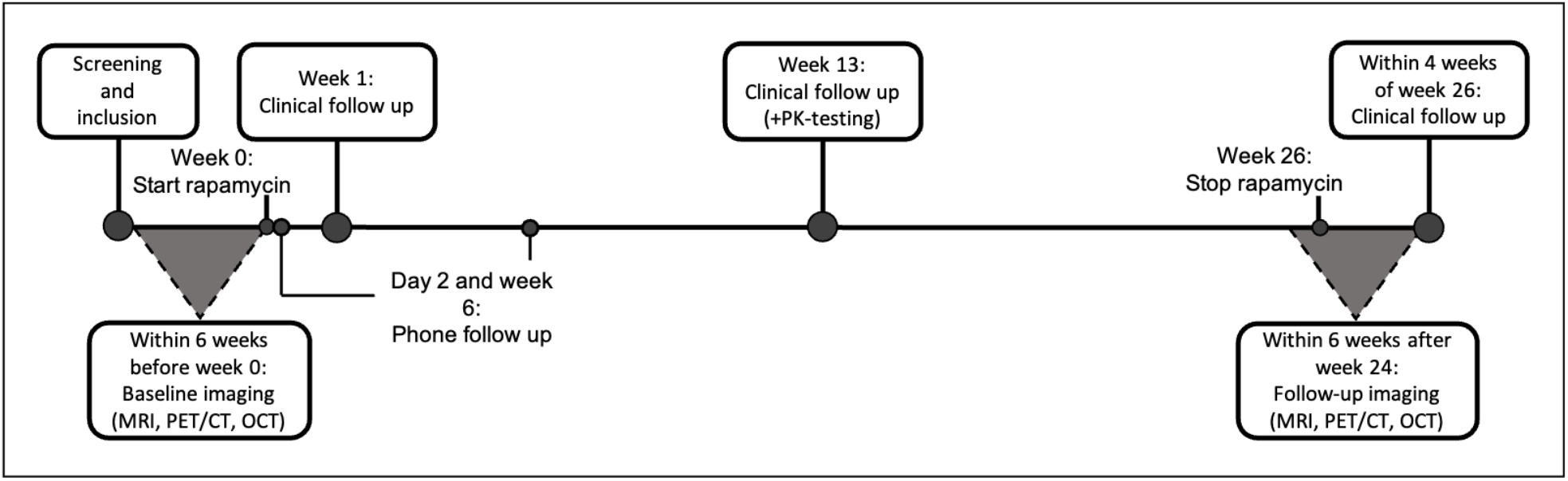
Study timeline for each participant.

### Participants

The study will enrol patients with early-stage AD, defined as fulfilling criteria for Alzheimer’s clinical syndrome, with either mild cognitive impairment (MCI) or mild dementia of the Alzheimer’s type, according to the NIA-AA (National Institute of Aging-Alzheimer’s Association) 2018 criteria (42) (see Box 1 for specific study eligibility criteria).

### Study drug

Rapamycin was approved in 1999 in the USA and in 2001 in Europe as an immunosuppressive drug to prevent organ rejection in renal transplantation (44). The drug and structurally analogues compounds (known as “rapalogs”), such as everolimus, have been approved for the treatment of several solid tumours (45,46), and is currently the only pharmacological option when treating tuberous sclerosis complex (TSC) (47). Rapamycin exerts its effect by inhibiting the intracellular protein kinase mTOR, which stands for “mechanistic target of rapamycin”. mTOR has been shown to be central in the regulation of several important functions in mammalian cells, such as cell growth and proliferation, protein synthesis, and autophagy (48).

The bioavailability of orally administered rapamycin is low (approximately 15%) and highly variable (SD = 9%). The drug is metabolized in the liver, primarily by CYP3A4, with a terminal half-life of 62 hours, though also here with large interindividual variability (SD = 16 hours) (43).

#### Adverse events, mitigation strategies and dosing

The side effect profile of rapamycin is well known, from a large number of clinical trials, and from long clinical use. The treatment is generally well tolerated, but common side effects, as described in the product information (44), are; stomatitis, diarrhea, and nausea. Changes in clinical laboratory values observed during rapamycin treatment include increased blood levels of cholesterol and triglycerides, and bone marrow depression manifesting as thrombocytopenia and anemia. The incidence of bacterial infections has been reported as increased in cancer patients treated with rapamycin, along with reports of cases of non-infectious pneumonitis (45).

Notably, the data on side effects is based on the use of rapamycin following organ transplantation, where the drug is commonly used together with other immunosuppressants. In the ERAP trial, we plan to deviate from the standard dosing of rapamycin in two ways. Typically, when used as an immunosuppressant, rapamycin is administered orally at a daily dose of 2 mg or above (44). We will instead administer an overall lower dose but in an intermittent fashion; a weekly oral dose of 7 mg. This change is aimed at reducing the risk of adverse events. The rationale behind this is that positive effects of rapamycin are hypothesized to be caused by inhibition of the mTOR1 complex, while many of the side effects are hypothesized to be due to inhibition of the mTOR2 complex. While mTOR1 is sensitive to acute dosing treatment, mTOR2 requires sustained exposure of the drug in order to be effectively inhibited (46).

Patients will be monitored for side effects during the study, including collection of blood samples at follow-up visits (see Table 1). These samples will be analyzed for standard clinical measures, including parameters that are known to be affected by rapamycin: complete blood count with differential and platelet count, sodium, potassium, chloride, albumin, creatinine, bilirubin, alkaline phosphatase, alanine aminotransferase, aspartate aminotransferase, gamma-glutamyl transferase, glucose, cholesterol, triglycerides, calcium, phosphorus, and creatine phosphokinase.

**Table 1.**
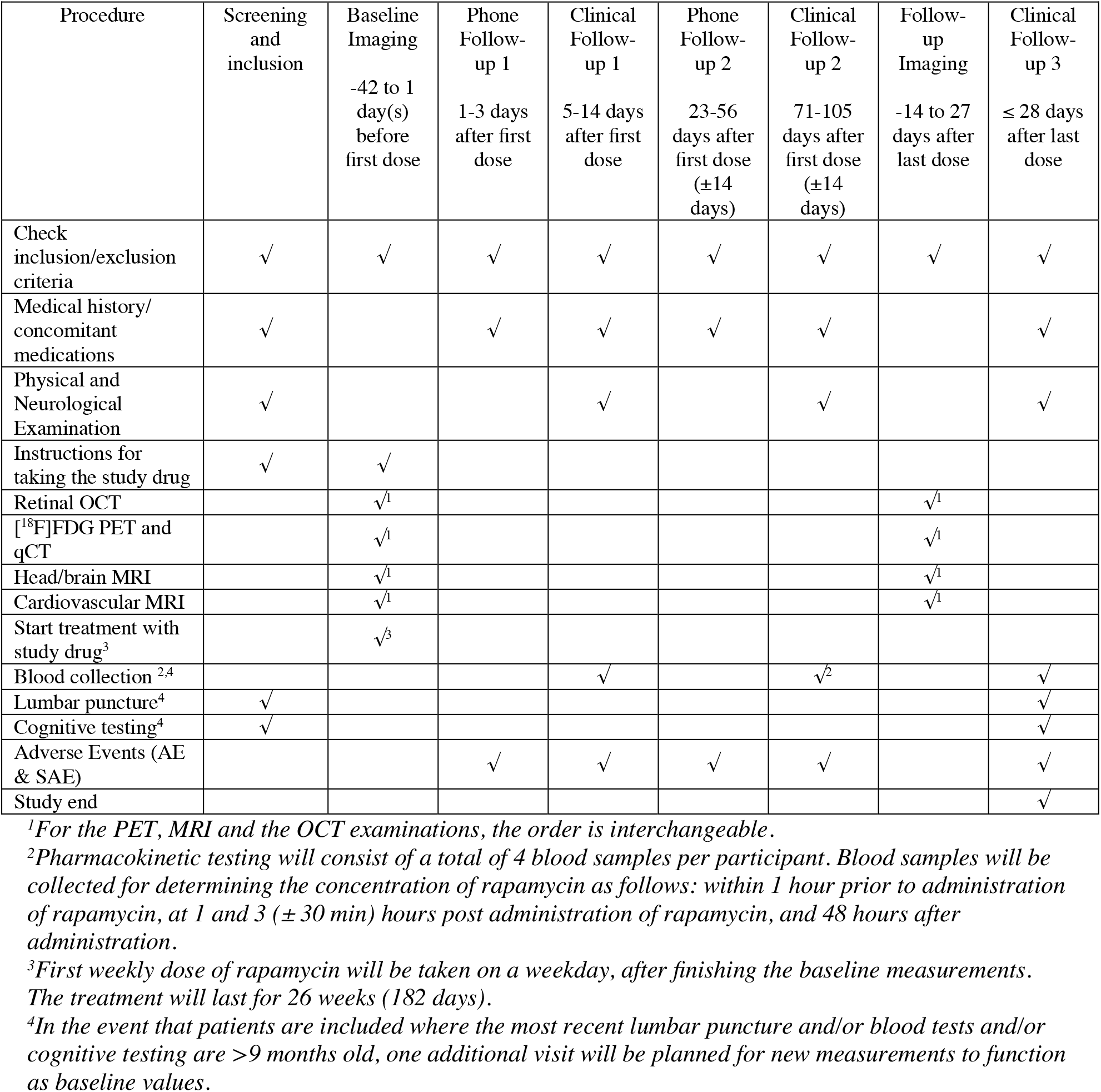
Visits, follow-ups and their corresponding assessments/examinaCons.

#### Blood-brain-barrier passage

It has not been thoroughly explored to which degree rapamycin passes the blood-brain-barrier (BBB) in humans. Rapamycin is a large molecule (molecular weight 914.2) and a substrate, albeit with low affinity, for the efflux pump P-glycoprotein (47). Drugs with these properties are often considered unlikely to pass from intestine to blood and bind an intracellular target (48). It is however known, from long clinical use, that oral treatment with rapamycin in humans leads to intracellular mTOR inhibition. The passage through cell walls, likely facilitated by high lipophilicity (logP estimated to be 4.3), supports BBB passage despite the large size of the molecule.

Following oral dosing, detectable rapamycin concentration in the brain of rodents has been established (49,50), and a large number of studies show clear effects in the central nervous system of animals (7). Support for cerebral target engagement (*i*.*e*. mTOR inhibition) in humans can be found from the use of rapamycin as a first-line treatment for the cerebral manifestations of TSC (51). TSC is a genetic disorder that activates the mTOR pathway, leading to the growth of benign tumors in organs, including the brain. Inhibition of mTOR with rapamycin analogues is the only approved pharmacological treatment of the disease, and the only feasible mechanism of action is mTOR inhibition in cells behind the BBB.

### Visits and data collection

Table 1 and Supplementary Information [Additional file 1] outlines the study visits, follow-ups, and corresponding assessments. In brief, participants will be invited to a first screening visit together with a study partner. During the visit the study will be explained in detail and written informed consent will be obtained. Basic clinical and demographic information will be collected, and the study eligibility criteria will be assessed (see Box 1).

Before initiating of study treatment, the following baseline examinations will be performed: [^18^F]Fluorodeoxyglucose ([^18^F]FDG) PET/CT imaging, brain and head MR imaging, cardiological MR imaging, retinal optical coherence tomography, lumbar puncture for collection of a CSF sample, as well as neuropsychological testing and physical aptitude. At the end of the treatment period the same set of follow-up examinations will be performed.

During the treatment period, participants will attend three clinical follow-up visits. At every visit, information on side effects will be collected. During the second clinical follow-up, blood will be collected at four time points over 48 hours to study the drug’s pharmacokinetic properties (just before and 1,3, and 48 hours after intake of the weekly dose). At the third clinical follow-up visit, which will occur after the final dose of the study drug, neuropsychological cognitive tests will be performed and a CSF sample will be collected. Additionally, each participant will have at least two scheduled phone calls during the study to assess adverse events or changes in concomitant medications/supplements.

### Objectives and endpoints

Table 2 presents the study objectives along with their respective outcomes and endpoints.

**Table 2.**
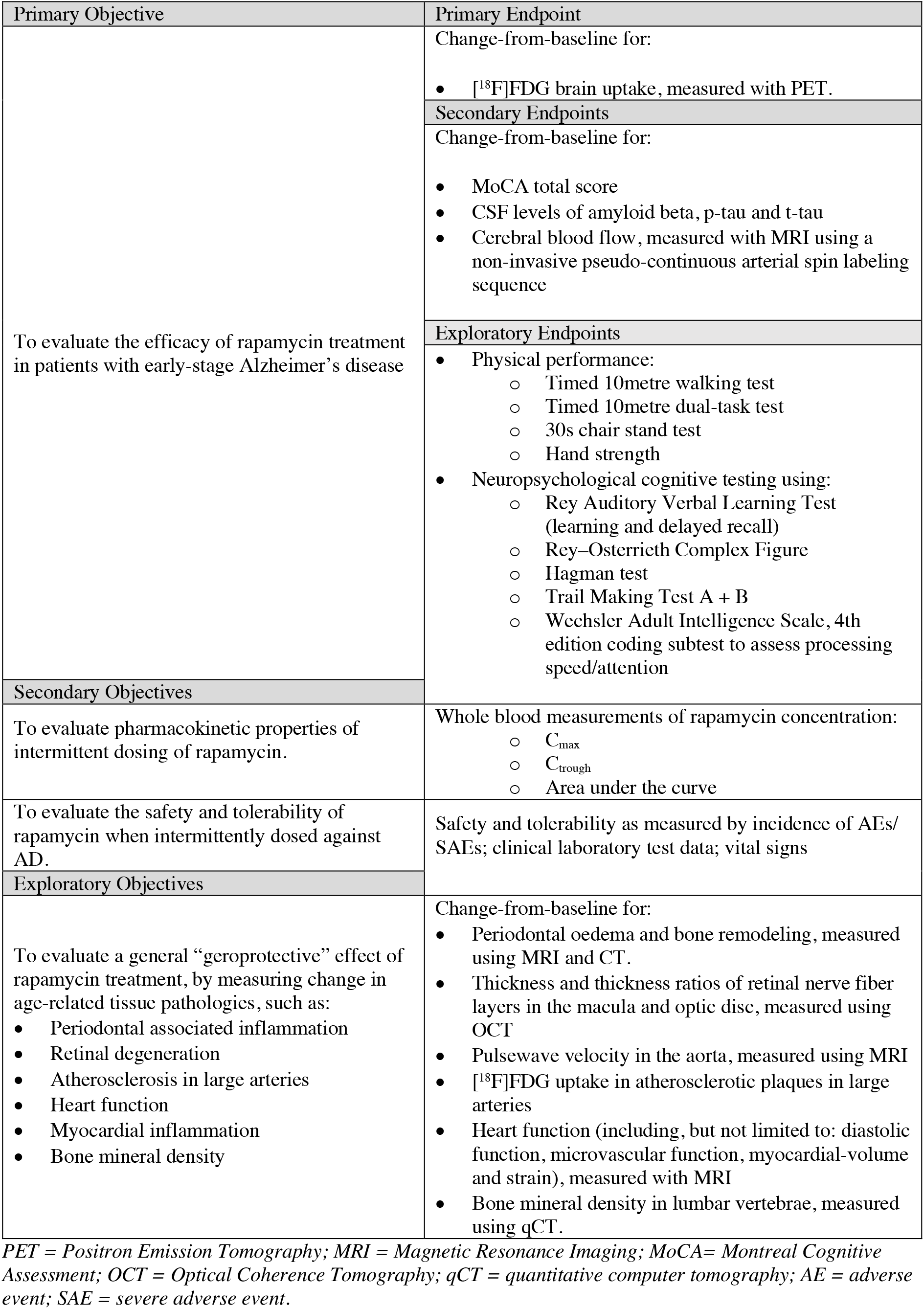
Study objecCves and endpoints.

#### Primary objective

The primary objective of ERAP is to evaluate the effect of rapamycin on the progression of early-stage AD. The primary endpoint will be the change in [^18^F]FDG PET uptake in the cerebral grey matter between baseline and the end of the study. Multiple studies have demonstrated that cerebral glucose metabolism, as assessed using [^18^F]FDG PET, declines progressively with normal aging and is further impaired in AD (52,53). Consequently, brain [^18^F]FDG uptake is commonly utilized as a diagnostic tool for AD and has served as a biomarker for disease progression when assessing the effectiveness of potential AD treatments (54).

Secondary endpoints for assessing treatment efficacy will be change between baseline and end-of-study in cerebral grey matter perfusion (blood-flow) measured using MRI and a pseudo-continuous arterial spin-labelling sequence; CSF levels of amyloid beta 42, phosphorylated tau and total tau, as well as change in the neuropsychological test the Montreal Cognitive Assessment (MoCA) total score.

#### Secondary objectives

The safety and tolerability of intermittently dosed rapamycin in early-stage AD will be assessed. We will monitor and record the incidence of treatment-emergent adverse events (AE), severe adverse events (SAE) by clinical follow-up examinations, where vital signs and blood tests will be assessed (see Supplementary Information 2 and 3 [Additional file 1]).

The pharmacokinetic profile of rapamycin has not been thoroughly studied in the setting of an intermittent dosing scheme. As a secondary objective, we will assess the differences in whole blood concentration of the study drug among individuals by comparing peak (C_max_), trough (C_trough_), and area-under-the-curve (AUC) concentrations. This will also allow us to assess if any potential differences in the treatment effect are associated with drug whole blood concentration among participants.

#### Exploratory objectives

An exploratory objective of this study is to quantify changes in multiple age-related tissue pathologies before and after treatment with rapamycin, using a set of imaging techniques (see Table 2). If a beneficial effect on multiple such pathologies can be shown, it will lend support to the hypothesis that the study drug has a general geroprotective effect in humans.

Exploratory outcomes will consist of assessments of change between baseline and end-of-study imaging outcomes including retinal nerve fibre layer thickness, periodontal oedema, arterial stiffness and [^18^F]FDG uptake in arterial plaques, cardiac diastolic function, myocardial strain, cardiac microvascular function as well as bone mineral density.

### Adverse events

Safety and tolerability will be assessed through monitoring and recording of all adverse events and serious adverse events. Clinically significant deviations in vital signs, laboratory evaluations, and physical examinations will be considered as adverse events and will be followed up. To the extent that it is possible, all adverse events will be described by their severity grade, duration and relationship to the study drug.

### Statistics

Based on the relatively low variability in long-term test-retest data of [^18^F]FDG in humans (55,56), a sample size of N = 15 is estimated to be sufficient to detect a 5% change in cerebral grey matter metabolic rate at 80% power with a significance level of 0.05. Such a hypothesized effect size is considered feasible given previous trials of AD using [^18^F]FDG as an outcome measure (57,58).

The change in estimated metabolic rate of grey matter [^18^F]FDG between baseline and follow-up imaging will be assessed using a paired t-test. Additionally, grey matter differences in standardized uptake value ratios, using the cerebellum as a pseudo-reference region (denominator), will be evaluated as a complementary outcome measure of [^18^F]FDG uptake. The level of significance will be set at 0.05.

Paired t-tests will also be used to assess differences between baseline and end-of-study secondary outcome measures. We will also explore if pharmacokinetics parameters are correlated with 1) each other, 2) side effect burden, 3) treatment effect using linear models.

### Ethical and Regulatory Considerations

The study will be conducted in accordance with the Declaration of Helsinki’ and the International Conference on Harmonisation for Good Clinical Practice (ICH GCP E6). The study protocol and relevant documents were approved by the Swedish Medical Products Agency (Läkemedelsverket, number: 5.1-2023-8283), and the Swedish Ethical Review Authority (Etikprövningsmyndigheten, number: 2023-03075-02 and 2023-00611-01), EudraCT number: 2023-000127-36. The trial has been registered at ClinicalTrials.gov (NCT06022068, first release August 30, 2023). Prior to study enrolment, informed consent will be obtained from each participant and their study partner.

## DISCUSSION

The ERAP trial is a phase IIa, one-arm, open-label, single-centre study designed to investigate the potential of the drug rapamycin to be repurposed as a treatment for early-stage AD. Repurposing an approved drug for a new indication has the potential to substantially reduce the cost and time of drug development (59). In the field of AD treatment research, 37% of candidate agents in the pipeline are repurposed drugs (60).

### Possible mechanisms of action

Pre-clinical data suggests that rapamycin may be an effective drug for treating neurodegenerative disorders (6,7). Several non-mutually exclusive mechanisms have been hypothesized to underlie this putative effect:

1. *Autophagy Regulation:* Inhibition of mTOR is known to upregulate cellular macro-autophagy (9). Deteriorating autophagy and increased mTOR activity have been observed in normal aging and in the progression of AD (61,62). Autophagy plays a central role in clearing intracellular toxic aggregate-prone proteins. Stimulation of autophagy by rapamycin could facilitate intercellular clearance of misfolded proteins central to the pathophysiology of AD.
2. *Vasculoprotection*: Reduced cerebral perfusion and compromised integrity of the blood brain barrier (BBB) have been suggested as drivers behind AD pathology (63,64), supported by observations of cerebrovascular dysfunction as one of the earliest detectable changes in AD patients (65). Rapamycin has been shown to improve cerebral perfusion and BBB integrity in rodent models of AD, supporting the notion of the mTOR pathway as a potential target for brain vasculoprotection in AD (66).
3. *Immunomodulation*: A sustained activation of microglia and ensuing inflammation is a central feature of neurodegenerative disorders, including in AD (67). Rapamycin’s effect on immune function is complex; while its main clinical use has been as an immunosuppressant, it has also been shown to augment immunity to certain pathogens (68), and improve response to influenza vaccination in elderly individuals (69). Beneficial immunomodulatory effects could be driven by an increase in T-regulatory (Treg) cell function. Tregs might play an important role in the treatment of AD by suppressing microglia-mediated inflammation (70). In line with this, a reduction in inflammatory CNS markers has been shown following rapamycin treatment (71), suggesting that this could be a potential mechanism for a treatment effect on AD.

### Assessment of general geroprotective properties

In addition to its potential as a treatment for AD, rapamycin has also been hypothesized to have a general geroprotective effect by slowing multiple age-related processes in the human body. In the ERAP trial, we aim to collect data on a wide range of age-related pathological processes using imaging techniques such as PET, MRI, CT, and retinal OCT. If positive changes are observed in multiple outcomes reflecting various age-related pathologies in different organs and tissues, it would support the hypothesis that rapamycin has a general geroprotective effect. The logistics of collecting and quantifying the listed exploratory imaging outcomes in Table 2 are facilitated by the fact that participants are already undergoing whole-body PET/CT examinations and MRI procedures for the trial’s primary and secondary endpoints. Adding sequences to quantify potential changes in additional pathologies can therefore be done with acceptable levels of additional discomfort and/or radiation exposure to participants.

### Limitations

The main limitations of this study are the absence of a control group, the small sample size and short trial duration. Without a control group, detecting any potential inhibition of AD progression is not possible, and the current design relies on an increase in cerebral glucose metabolism in a relatively short time to demonstrate a positive treatment effect. However, ERAP is a phase IIa trial aimed at generating exploratory data on the effect of rapamycin on AD, and assessing the feasibility of conducting a larger, longer and controlled clinical trial using imaging outcomes as endpoints in the future.

## CONCLUSIONS

The study will measure a set of AD biomarkers before and after a 6-month dosing scheme, with the primary endpoint being change in [^18^F]FDG PET uptake in the cerebral grey matter, a well-established diagnostic and prognostic biomarker of AD disease progression. The findings from this repurposing effort of rapamycin can provide evidence of a novel treatment alternative for Alzheimer’s disease and form the basis for larger controlled phase IIb or III trials. This study will also investigate the potential general geroprotective effects of rapamycin on various age-related pathologies in the human body.

## Data Availability

The data collected as part of the trial will be shared in accordance with institutional regulations.

## DECLARATIONS

### Ethical Approval and consent to participate

The study protocol and relevant documents were approved following review by the Swedish Medical Products Agency (Läkemedelsverket, number: 5.1-2023-8283), and the Swedish Ethical Review Authority (Etikprövningsmyndigheten, number: 2023-03075-02 and 2023-00611-01), EudraCT number: 2023-000127-36. The trial has been registered at ClinicalTrials.gov (NCT06022068, first release August 30, 2023). Prior to study enrolment, informed consent will be obtained from each participant and their study partner.

### Consent for publication

Not applicable

### Availability of data and materials

Not applicable

### Competing Interest

The authors declare that they have no competing interests.

### Funding

The study is supported by a Longevity Impetus grant from the Norn Group, Åhlén Stiftelsen, Demensfonden, The Swedish Society of Medicine (SLS), Loo and Hans Osterman Stiftelse, Stiftelsen för Ålderssjukdomar Karolinska Institutet, Stiftelsen för Gamla Tjänarinnor, Tore Nilssons Stiftelse för Medicinsk Forskning, Stiftelsen Stockholms Sjukhem, Region Stockholm (ALF grant), The Swedish Brain Foundation (PS2021-0012), and KI CIMED. None of the funding bodies had any role in the design of the study or in writing the manuscript.

### Authors’ Contribution

All authors contributed to the development and/or the preparation of the study protocol; PPS and JS are responsible for conception and overall design of the study and obtained funding. PPS and JS prepared the first draft of the manuscript; all authors reviewed and approved the final version of the manuscript.

## Acknowledgments

We would like to thank Edvin Johansson, Martin Schain and Lars Johansson from Antaros Medical for their assistance with the design of MRI and PET imaging protocols. We would like to thank Lars Farde for his mentorship and feedback on the study design.

## Box 1.

**Eligibility Criteria**

**STUDY ELIGIBILITY CRITERIA**

*Inclusion criteria*

1. Age 55-80 years.
2. Has an available “study partner” that can accompany the participant to planned visits.
3. Has a clinical diagnosis of MCI (mild cognitive impairment), or dementia of Alzheimer’s type, and:
  - At inclusion, the participant meets the criteria for “Alzheimer’s clinical syndrome”, MCI or mild dementia of the Alzheimer’s type, according to the NIA-AA (National Institute of Aging-Alzheimer’s Association) 2018 criteria (42).
  - At inclusion, the participant is amyloid positive, established with either amyloid PET imaging, or a CSF beta amyloid 1-42 assay, or a CSF beta amyloid 1-42/ beta amyloid 1-40 assay.

3.1 For participants with dementia, the disease should be in an early stage, operationalized as:
  - Having stage 4 mild dementia or lower, according to the NIA-AA 2018 clinical staging criteria (42).
  - Having a clinical Dementia Rating Scale (CDR) global score of 1 or lower.
  - Having a Montreal Cognitive Assessment (MoCA) score of ≥ 18 or a Rey Auditory Verbal Learning Test (RAVLT) > 4 words after 30 minutes.
3.2 For participants with a diagnosis of MCI, a cognitive deficit with >-1SD in at least one of the following cognitive tests: Wechsler Adult Intelligence Scale subtest to assess processing speed/attention, Rey Auditory Verbal Learning Test (learning and delayed recall) or Rey Complex Figure Test.

4. Is proficient in the Swedish language.
5. Has a normal or clinically acceptable medical history, physical examination, and vital signs.
6. For female participants, the participant has no childbearing potential, meaning that she is surgically sterile or post-menopausal, or a negative pregnancy test following a menstrual period AND use of an acceptable effective contraceptive measure, which must be continued at least 12 weeks after stopping the study drug.

*Exclusion criteria*

1. Has a history of any major disease that may interfere with safe engagement in the intervention (especially severe liver or kidney disease, or uncontrolled diabetes).
2. Has a history of a major neurological disorder, central nervous system infarct, infection or focal lesions of clinical significance on MRI scans.
3. There is evidence of a clinically relevant or unstable psychiatric disorder, based on Diagnostic and Statistical Manual of Mental Disorders (DSM-5) criteria, including schizophrenia or other psychotic disorder, or bipolar disorder.
4. Fulfill any contraindication for the use of rapamycin, including (but not restricted to):
  - Current or planned medication with a strong inhibitor of CYP3A4 or P-gp.
  - Current or planned medication with a strong inducer of CYP3A4 or P-gp.
  - Other current medications with known serious interaction risks with rapamycin.
  - Known allergy or hypersensitivity to rapamycin.
5. Has significant obesity, per the investigator’s judgement.
6. Has untreated hyperlipidemia that is clinically significant, per the investigator’s judgement.
7. Has undergone treatment with immunosuppressive medications within the last 90 days, or treatment with chemotherapeutic agents for malignancy within the last 3 years.
8. Has had major surgery within 3 months prior to the planned start of rapamycin treatment, or has major surgery planned during the period of the trial.
9. Has used experimental medications for AD or any other investigational medication or device within the last 60 days of inclusion.
  - Participants who have been involved in a monoclonal antibody study are excluded unless it is known that they were receiving placebo in that trial.

## Notes

### Competing Interest Statement

The authors have declared no competing interest.

### Clinical Trial

NCT06022068

### Funding Statement

The study is funded by a Longevity Impetus grant from the Norn Group, Åhlén Stiftelsen, Demensfonden, The Swedish Society of Medicine (SLS), Loo and Hans Osterman Stiftelse, Stiftelsen för Ålderssjukdomar Karolinska Institutet, Stiftelsen för Gamla Tjänarinnor, Tore Nilssons Stiftelse för Medicinsk Forskning, Stiftelsen Stockholms Sjukhem, Region Stockholm (ALF grant) and KI CIMED. None of the funding bodies had any role in the design of the study and in writing the manuscript.

### Author Declarations

The Swedish Ethical Review Authority (Etikprövningsmyndigheten, number: 2023-03075-02 and 2023-00611-01) gave ethical approval for this work.

### Summary of Updates

Title revised, abstract updated, funding information added.

