## Supplementary Information for "Evaluating the effect of rapamycin treatment in Alzheimer's Disease and aging using *in vivo* imaging: the ERAP phase IIa clinical study protocol"

**SUPPLEMENTARY INFORMATION 1:** Information, data and outcomes collected at visits and follow-ups.

*Screening*

- Relevant medical history and coexisting disease.
- Use of all medication, vitamins and supplements during the study and within three months of entry.
- Complete physical examination including neurological examination; pulse, blood pressure, height, and weight.
- Timed 10-meter walking test.
- Timed 10-meter dual-task test.
- 30s chair stand test.
- Participant demographics and medical history.
  - Date of birth.
  - Sex.
  - Education.
  - Date of AD or MCI diagnosis.
  - Family history of AD (first degree relatives).
  - Smoking habits.
  - Alcohol consumption.
- Neuropsychological cognitive tests. If there exist test scores for the participant that is not older than 9 months, this data will be used instead.
  - Rey Auditory Verbal Learning Test (learning and delayed recall).
  - Rey–Osterrieth Complex Figure (ROCF).
  - Hagman test.
  - Trail Making Test (TMT) A + B.
  - Montreal Cognitive Assessment (MoCA).
- CSF sample. If there exists a CSF sample for the participant that is not older than 9 months, this data will be used instead.

*Baseline and follow-up imaging*

- Whole-body positron emission tomography (PET) [$^{18}$F]FDG scan and quantitative computer tomography (qCT) scan on the integrated PET/CT system.

- Head/brain magnetic resonance imaging scan (MRI).
- Cardiac MRI scan.
- Retinal optical coherence tomography (OCT) scan.

*Clinical follow up 1, 2 and 3*

- Inclusion and exclusion criteria.
- Adverse events.
- Changes in use of concomitant medication and supplements.
- Treatment compliance (participants will be instructed to bring their dosette box for inspection).
- Physical examination, including:
    - Pulse, blood pressure, weight.
    - Timed 10-meter walking test.
    - Timed 10-meter dual-task test.
    - 30s chair stand test.
- A venous blood sample will be collected and analyzed for blood values in connection with the visits.

Additionally, at clinical follow up 2 blood data will be collected for pharmacokinetic analysis. The visit will be booked for the day of the weekly dosing and will be scheduled between the third and last dose of the study drug.

- Venous blood test will be drawn at three time points while at the clinic (prior to taking the study drug, and at 1 and 3 hours post dosing), and once after 48h.

Additionally at clinical follow up 3 (last visit):

- CSF sample.
- Follow-up cognitive testing (see Screening for list).

*Phone follow-up*

- Inclusion and exclusion criteria.
- Adverse events.
- Changes in use of concomitant medication and supplements.

Following a phone follow-up, a physical visit can be booked if deemed necessary by the investigator.

**SUPPLEMENTARY INFORMATION 2:** Procedures for measurement of endpoints.

**Primary endpoint**

*PET/CT*

The metabolic rate of glucose (MRGlu) in central nervous system grey matter measured through [$^{18}$F]FDG PET will be the primary endpoint.

Participants will be invited to [$^{18}$F]FDG PET/CT scans at two occasions, within four weeks before, and by the end-of-treatment. Prior to the scan session, participants will be asked to fast for a minimum of four hours.

After pre-scan preparations, the participants will be asked to lay recumbent in the PET/CT system on top of a CT calibration cylinder. A qCT scan will be acquired over the L1-L3 vertebrae using the integrated CT module, to assess the bone mineral density in the trabecular and cortical bone compartments as well as muscle volume and density in the paravertebral muscles. The CT module will then be used to acquire an attenuation scan, which is necessary for the ensuing PET image reconstruction.

Prior to injection of [$^{18}$F]FDG radioligand the participant will receive a peripheral venous catheter in each arm. A blood sample will be drawn and glucose level will be quantified. The target dose for the injected radioactivity of [$^{18}$F]FDG will be 2 MBq/kg (i.e., 140 MBq for a 70kg participant, translating to an effective dose of 2.7 mSv). A sterile physiological phosphate buffer (pH 7.4) solution containing the radiotracer will be diluted with saline to the volume of 10 ml and then injected as a bolus during 10 seconds into a cannula inserted into an antecubital vein. The cannula is then immediately flushed with 10 ml saline.

For the [$^{18}$F]FDG scan, emission data will be acquired in list mode during up to 60 min. For the initial 10 min after injection, the field-of-view will be centered over the thorax to obtain a high temporal resolution dynamic imaging of the descending aorta. Following this, there will be a pause in scanning for 20min and then the field-of-view will be centered over the head and new emission data will be acquired in 5 min time frames for 30 minutes. At the 60 min mark after injection, a final emission acquisition will be performed over the thorax again for 20 minutes.

During the duration of the emission scans, 5 venous blood samples will be collected from the left arm (first sample at 20 min, last sample at 90 min after injection), and the radioactivity of blood will be measured.

PET images will then be reconstructed into frames and motion corrected to the acquired CT. Image preprocessing and modelling will be done in a blinded fashion, where the analyst does not know which scan is the baseline and which scan is the follow-up.

Acquired structural brain MRI-images (see below) will be co-registered to PET-images and a grey matter mask will be delineated using automated software. A manual delineation of the descending aorta will then be done on the CT image. The decay corrected radioactivity uptake in frames will be extracted from these masks, and an input function will be created using the time-activity curve from the delineated descending aorta mask concatenated with the radioactivity in the five venous samples. This input function will then be used together with the radioactivity time activity curve in the grey matter mask to quantify the net transfer rate ($K_i$), using the Patlak Graphical Method (“Patlak plot”) (1). This estimate will then be corrected for participant blood glucose levels to derive an estimate of the MRGlu.

**Secondary and exploratory endpoints**

*MRI examination*

Before treatment initiation participants will perform two separate MRI examinations; one head/brain scan and one cardiac scan. At the end of the treatment period participants will repeat the same set of MRI examinations again.

Several different tissues will be examined using a set of different protocols:

- A T1-weighted structural brain image acquired using a Magnetization Prepared - Rapid Gradient Echo sequence, for structural and volumetric information.
- Pseudo-continuous arterial spin labeling sequence for assessing change in grey matter perfusion (cerebral blood flow).
- A T2-weighted image over the alveolar bone, including teeth and periodontal tissues acquired using the Short Tau Inversion Recovery sequence for assessing periodontal oedema in the upper jaw.
- An aortic pulsewave velocity estimation acquired using through-plane phase-contrast MRI at ascending and descending aortic sections, with time-to-peak estimates used to derive aortic stiffness.

- Cardiac function and structure assessed via cardiac MRI including parameters such as:
  - Ejection Fraction
  - Strain
  - Ventricular and atrial volumes
  - Ventricular mass
  - Extracellular volume
  - Myocardial perfusion

*OCT*

Imaging of the optic disc and macular cubes will be done by an OCT scan. For the optic disc cube, quantification will be carried out of the average total retinal nerve-fiber layers thickness, as well as the specific thickness of the four standard quadrats of both oculus dexter and sinister. For the macular cube, quantification will be carried out of the average ganglion cell layer thickness -+ the inner plexiform layer thickness, average thickness of the six sectors as well as thickness ratio between the highest and lowest sector on both oculus dexter and sinister.

*Laboratory Assessments (Blood and CSF samples)*

All biosamples (blood and CSF) will be analyzed at clinical laboratories at Karolinska University hospital.

A lumbar puncture will be performed at screening (unless a <9-month-old sample already exists in case data from such sample will be used instead) and at the last clinical follow-up. Up to 20 mL CSF will be sampled. The following variables will be quantified and used as secondary endpoints: amyloid beta (40 and 42), P-tau and T-tau.

Blood tests for safety assessment and assessment of baseline and follow up variables will be collected on up to 4 occasions. The following parameters will be quantified and assessed at follow up 1 and 2.

- Hematology: Complete blood count with differential and platelet count.
- Blood chemistry: Sodium, potassium, chloride, albumin, creatinine, bilirubin (total and direct), alkaline phosphatase, alanine aminotransferase, aspartate aminotransferase, gamma-glutamyl transferase, glucose, cholesterol, triglycerides, calcium, phosphorus, creatine phosphokinase.

At screening and clinical follow up 3 (i.e., end of treatment) a more comprehensive panel of blood tests used during the clinical work up at the memory clinic will be analyzed.

Part of the samples (both blood and CSF) will be stored in a biobank and might be retrieved and used for further exploratory analyses of the research questions outlined in this protocol.

*Pharmacokinetic assessments*

As a secondary objective we will examine the pharmacokinetic properties of intermittently administered rapamycin. The endpoint variables will be the whole blood concentration of the study drug at timepoints for expected through ($C_{trough}$), and peak ($C_{max}$) concentrations, and at approximately 48h post-dosing. During the clinical follow-up 2, which will be scheduled to the dosing day of the week 13 +- 2 weeks, four blood samples will hence be collected for determining the concentration of rapamycin as follows:

- Within 1 hour prior to administration of sirolimus,
- At 1 and 3 hours post administration of sirolimus,
- 48h post-dose

**SUPPLEMENTARY INFORMATION 3:** Monitoring and handling of adverse events.

Confirmed abnormal laboratory results (after repeat) will be assessed for clinical significance and reported as AEs if necessary. Laboratory abnormalities assessed as clinically significant have to be reported as an AE only if they meet at least one of the following conditions:

- The abnormality suggests a disease and/or organ toxicity AND this abnormality was not present at the screening visit, or is assessed as having evolved since the screening visit.
- The abnormality results in discontinuation of the study drug.
- The abnormality requires medical intervention or concomitant therapy.

The following strategies will be employed with regards to specific side effects:

- Increase in fasting glucose: if a fasting glucose above 6 mmol/l is detected and sustained after a second measurement, an oral glucose tolerance test will be performed. If needed (as per the investigators judgment), the results of this test will be discussed with the endocrine consultant at Karolinska University hospital and/or the participants primary care physician. One of the following ways forward will be decided on together with the study participant: 1) no change in treatment and continued monitoring; 2) reduction of dose of rapamycin (or increased time between doses), followed by new tests; 3) discontinuation of rapamycin; 4) temporary treatment with anti-diabetic medication during the remaining study period.

- Dyslipidemia: if the lipid profile shows a clinically significant change (as per the PIs assessment) compared with baseline. If needed (as per the investigators judgment) the results of this test will be discussed with the cardiology consult at Karolinska University hospital and/or the participant's primary care physician. One of the following ways forward will be decided on together with the study participant: 1) no change in treatment and continued monitoring; 2) reduction of dose of rapamycin (or increased time between doses), followed by new tests; 3) discontinuation of rapamycin; 4) temporary treatment with a statin during the remaining study period.
- Bone marrow suppression: blood panel showing a clinically significant (as per the PIs assessment) thrombocytopenia, anemia or leucopenia. If needed (as per the investigators judgment) these results will be discussed with the hematology consult at Karolinska University hospital and/or the participant's primary care physician. One of the following ways forward will be decided on together with the study participant: 1) no change in treatment and continued monitoring; 2) reduction of dose of rapamycin (or increased time between doses), followed by new tests; 3) discontinuation of rapamycin.
- Electrolyte imbalance: electrolyte panel showing a clinically significant (as per the PIs assessment) change compared with baseline measurements. If needed (as per the investigator's judgment) these results will be discussed with the internal medicine consult at Karolinska University hospital and/or the participant's primary care physician. One of the following ways forward will be decided on together with the study participant: 1) no change in treatment and continued monitoring; 2) reduction of dose of rapamycin (or increased time between doses), followed by new tests; 3) discontinuation of rapamycin.
- Immunosuppression: study participants will be instructed to temporarily stop the rapamycin treatment and seek medical attention if they experience fever chills, measure a body temperature above 38°C, or detect other signs of an infection. Participants should thereafter contact a study representative as soon as possible and the PI will assess whether the rapamycin treatment can be resumed.
- If a subject develops clinical signs indicating pneumonitis, rapamycin treatment will be temporarily stopped and a diagnostic work up, including chest radiology, will be ordered. If needed (as per the investigators judgment) the results will be discussed with the pulmonologist consultant at Karolinska University Hospital before: 1) restart treatment with continued monitoring; 2) restart treatment with reduced dose of rapamycin (or increased time between doses); 3) discontinuation of rapamycin.
